# Pigeon-Guano-Contaminated Environments in Blantyre, Southern Malawi, are Reservoirs of Medically Important Fungi

**DOI:** 10.64898/2026.05.26.26354139

**Authors:** Bashir Joey Merico, Petros Chigwechokha, Paul Alubino, Gama Petulo Bandawe

## Abstract

Close to 50% of all bird species are reservoirs of potentially pathogenic fungi, including species listed as priorities by the World Health Organization. In Malawi, data on the diversity, pathogenic potential, and avian ecological sources of medically important yeasts are scarce. A descriptive cross-sectional study was conducted to characterise medically important yeast recovered from environments contaminated with pigeon guano in Blantyre, Southern Malawi. A total of 20 pigeon guano samples were collected from 4 peri-urban areas, which yielded 71 yeast isolates. Only 54 isolates were successfully identified after VITEK MS analysis. More than 80% of the identified environmental isolates belong to fungal species listed as priority, with *Pichia kudriavzevii* (39%) and *Candida orthopsilosis* (30%) the most common species. Phenotypic comparisons of virulence traits: phospholipase activity, haemolysin activity, catalase, urease, biofilm formation, thermotolerance, and adhesion properties were made with 21 clinical yeast isolates from referral laboratories. *Candida parapsilosis sensu stricto* (29%) and *Candida albicans (24%)* constituted a majority of the clinical yeast isolates. Most of the environmental isolates exhibited virulence traits comparable to or stronger than those of clinical isolates, indicating an infection risk to humans and the urgent need for integrated, One Health-focused surveillance at bird-human interfaces.

## 1.0 Introduction

Each year, an estimated 300 million people worldwide suffer from invasive or systemic fungal infections, and about 1.65 million people succumb to them (1), exceeding deaths from severe diseases such as malaria, tuberculosis, and some cancers, and incurring a substantial public health cost (2). In the United States of America, for example, healthcare costs related to fungal infections were estimated to be more than $48 billion in 2017 (3). The situation in Africa is less well understood, primarily due to inadequate diagnostic facilities for fungal infections and the resulting paucity of accurate health data (4,5). Nevertheless, a very high prevalence of fungal infections has been reported in some African countries (6). An estimated 3 million cases of fungal infections occur each year in South Africa, and over 11.8% of the Nigerian population suffer from invasive fungal infections each year (7,8). In Malawi, it was estimated that about 1,338,523 (7.54%) people were affected by a serious fungal infection in the year 2018 (9).

Fungal infections are more invasive in immunocompromised hosts, causing considerable morbidity and mortality (10). Nearly 70% of all invasive fungal infections globally are caused by *Candida* species, followed by *Cryptococcus* species (20%) and *Aspergillus* species (10%) (11). In Malawi, among immunocompromised individuals, candidiasis is the most common recurrent infection; cryptococcal meningitis is the most common life-threatening infection; histoplasmosis is the second-leading cause of severe pulmonary infection after *Mycobacterium tuberculosis* (MTB); and pneumocystis pneumonia is the most life-threatening pneumonia (9). The introduction of azoles more than a decade ago has had a significant impact on curtailing invasive fungal infections; however, a dramatic increase in azole resistance has also been documented in recent years (11).

In 2022, the World Health Organization presented the first global systematic prioritisation of health-hazardous fungi (12). Under this prioritisation list, some notable species like *Cryptococcus neoformans* and *Candida albicans* were ranked as critical priority*, Histoplasma* species*, Candida tropicalis, Candida parapsilosis,* and *Nakaseomyces glabratus* as high priority, and *Pneumocystis* species, *Pichia kudriavzevii,* and *Cryptococcus gattii* as medium priority group (12,13). The systemic prioritisation was based on several criteria, among which were the extent of geographic distribution, the number of new cases per million population each year, and transmission or acquisition dynamics (12).

Animals often play a significant role in disease epidemiology (13,14). About 75% of all emerging and 61% of all infectious diseases are zoonotic (13,14). Birds cover long distances and can carry microorganisms across continents, making them important agents in zoonotic spread (13). Around 50% of all bird species are reservoirs and carriers of medically important fungi (15). Pigeons are common in urban areas as domestic and feral birds living close to humans (13). Their droppings (guano), are also known to be a reservoir for several pathogenic microorganisms, particularly bacteria such as *E. coli*, *Campylobacter* species, *Salmonella* species, and *Chlamydia* species (16). Despite the presence of several studies linking pigeons with other pathogenic species belonging to the genera *Candida*, *Histoplasma*, and *Aspergillus* (13,16,17), relatively few investigations have focussed on fungi outside of a handful of studies that have looked at some *Cryptococcus* species.

In Malawi, poultry, which includes pigeons, accounts for about 49% of all domesticated animals (18). Despite the high burden of fungal-associated infections in Malawi and a clearly established link between birds and fungal infections, there have been very few studies to assess the roles of birds in the transmission dynamics of medically important fungi and their pathogenic potential. Blantyre is among the districts in southern Malawi with a high HIV and fungal disease burden (9,19,20), and where pigeons are usually observed in urban areas close to human settlements. This makes it a critical setting for studying the potential of fungal pathogen transmission to humans via avian pathways. Therefore, this study was conducted to investigate the diversity and pathogenic potential of medically important fungal species in environments contaminated with guano from synanthropic pigeons at households in Blantyre.

## 2.0. Materials and Methods

### 2.1. Study Design and Area

A cross-sectional study was conducted between March 2025 and July 2025. Twenty households were identified based on proximity to locations where synanthropic pigeons are commonly observed, accessibility, and the willingness of the residents to permit sample collection. The households were located in 4 peri-urban areas in Blantyre (15°47′05″ S; 35°00′30″ E): Ndirande, Chilobwe, Mbayani, and Chemusa. These locales were selected as they represent areas of rapid urban sprawl, informal settlements, and had all been severely affected by recent impacts from natural disasters, which are some of the known drivers linked with the emergence, proliferation, and spread of fungal pathogens (21–23).

### 2.2. Ethical Approval

Before commencement of the study, ethical approval was sought from the Malawi University of Science and Technology Research Ethics Committee (MUSTREC), reference number: P.12/2024/373. MUSTREC is registered with the USA Office for Human Research Protections (OHRP) as an International IRB (IRB Number IRB00012588 FWA00029630). All participants/household owners involved in this study gave their formal informed consent verbally to participate. Initially, we clearly explained the study’s aims and the sampling procedure within the pigeon pen house to the household owners. Each household owner had the right to decline or participate without coercion or any undue incentives. All participation was voluntary, and owners could withdraw at any time without consequence. For the clinical isolates, approval and consent were granted by the laboratories responsible for each facility and the isolates were collected without any identifiable patient information.

### 2.3. Collection and Processing of Samples for Yeast Isolation

In this study, guano samples collected from the selected households are referred to as environmental samples; hence, isolates recovered from these samples are also referred to as environmental isolates. The guano samples were collected aseptically from each pigeon pen house using a multistage sampling approach. Subsequently, the samples from each cluster were pooled into a single sterile zip-lock bag, achieving a total mass of at least 50 grams. Pooling was performed to combine representative portions from each cluster of the pen house to enhance representativeness for culture. The samples were processed at the University of Malawi, Microbiology Laboratory, in accordance with previous studies (24,25). Processed samples were streaked onto Sabouraud Dextrose Agar (SDA) plates (MAST Group, Liverpool, United Kingdom) supplemented with 0.5% chloramphenicol. Each sample was inoculated in duplicate and incubated at 37°C and room temperature (25°C) for a period of one to seven days with daily visual inspection. This yielded 71 yeast isolates based on distinct colony morphologies.

### 2.4. Acquisition of Clinical Yeast Isolates

To understand the pathogenic potential of the environmental isolates, we sought to compare their diversity and some phenotypic virulence traits with those of human-associated yeast species. Therefore, we collected twenty-one (21) clinical yeast isolates from referral laboratories, i.e., the Zomba Central Hospital Laboratory (ZCHL), the Malawi Liverpool Welcome (MLW), and the Public Health Institute of Malawi. The isolates originated from candidemia cases, bloodstream infection, and urinary tract infection samples that had previously been processed at the facilities over the last 5 months, prior to commencement of the study.

### 2.5. Identification of Clinical and Environmental Yeast Isolates

Identification of all isolates was performed using the VITEK MS platform (BioMérieux, France) at the Central Veterinary Laboratory (CVL) in Lilongwe, Malawi, except for clinical isolates from PHIM identified at their facility with the same platform. Isolates from MLW and ZCHL were presumptively identified as *C. albicans* at their facilities using culture methods and later reassigned after VITEK MS analysis. At CVL, isolates were prepared per the National Microbiology Reference Laboratory VITEK MS protocol. Fresh, 24-hour *E. coli* ATCC 8739 served as control. A thin layer of fresh fungal isolates was applied in the VITEK MS slide sample circle in duplicate; 2 μL of formic acid was added to each circle to crystallize, then 1 μL of matrix was applied and allowed to dry before loading. Results were accepted when confidence exceeded 98%.

### 2.6. Determination of Virulence Factors

Virulence factors are what differentiate pathogenic organisms from the hundreds of species of non-pathogenic or commensal ones (26). Different species of fungi utilise several pathogenic mechanisms for survival (27). We evaluated multiple key virulence factors for the environmental and clinical yeast isolates. The temperature conditions remained constant across all tests: 37°C for thermotolerant species and room temperature (25°C) for thermosensitive species.

#### 2.6.1. Thermal tolerance

Only about 10% of the estimated 6 million fungal species are associated with human health, mainly due to their inability to bypass mammalian thermal barrier (3,28). Thermal tolerance was determined from the initial culture on SDA incubated at 37°C. All yeast isolates from which their colonies showed growth after incubation at 37°C were considered thermotolerant species. Species that were only isolated from culture plates incubated at 25°C were considered thermosensitive.

#### 2.6.2. Assessment of Biofilm Formation

Biofilm formation is associated with increased antifungal resistance and persistence of the pathogens on surfaces (27,29). Formation of biofilms was determined using the micro-plate technique as previously reported by others (30). Yeast colonies were suspended in 0.89% NaCl to match 0.5% McFarland. A 100 μL yeast suspension of each isolate was added to a 96-well plate (CytoOne, USA Scientific, USA) in triplicate and incubated for 1.5 hours with mild agitation. The first and last three wells served as controls. Following incubation, the wells were washed twice with 200 μL Phosphate Buffered Saline (PBS) to remove planktonic cells. Each well was filled with 150 μL Nutrient Broth (Sisco Research Laboratories, Mumbai, India) supplemented with 3% glucose and 0.5% chloramphenicol, then incubated for 60 hours to form biofilms. After two PBS washes and a 45-minute air-drying, 200 μL of 0.4% crystal violet was added for 45 minutes. Afterwards, the wells were washed, destained with ethanol, and 150 μL of the destained ethanol was read spectrophotometrically at a 570 nm wavelength with a microplate reader (Multiskan FC, Thermo Fisher Scientific, USA). Biofilm production was determined by averaging the Optical Densities (ODs) of each sample. In our study, isolates were classified into four categories based on their OD values: non-producers (ODs ≤ ODnc), weak producers (ODnc < ODs ≤ 2 × ODnc), moderate producers (2 × ODnc < ODs ≤ 4 × ODnc), and strong producers (ODs > 4 × ODnc).

#### 2.6.3. Adhesion to Biotic Surface

The successful establishment of infection by most pathogens is greatly dependent on the pathogen’s ability to adhere to host cells (31–33). The ability to adhere to biotic surfaces was tested using the human buccal epithelial cells (HBEC), based on a test procedure reported by others with a few modifications (30,34). The HBEC used in this study were harvested from a healthy individual. Initially, the test isolates were grown overnight to a stationary phase in nutrient broth (NB) supplemented with 3% glucose and 0.5% chloramphenicol. A standard yeast suspension was mixed gently with HBEC, at a ratio of at least 10 yeast cells per a HBEC and incubated in a rotary incubator for 1 hr. The yeast-HBEC suspension was washed four times with sterile PBS to remove unattached yeast cells. The preparations were examined as a direct wet preparation on an optical microscope under X25 objective lens (Brunel Microscope Limited, United Kingdom). In our study, the number of yeast cells attached to 50 HBEC was counted and interpreted as follows: weak if 0-10 yeast cells/50 HBEC, moderate if 11-25 yeast cells/50 HBEC, strong if 26-50 yeast cells/50 HBEC, and extremely strong if >50 yeast cells/50 HBEC.

#### 2.6.4. Adhesion to Abiotic Surface

The ability to adhere to abiotic surfaces was assessed using the biofilm formation assay performed in 96-well polystyrene micro-plates (28). Since the initial stages of biofilm formation involve the reversible and irreversible attachment of planktonic cells to surface materials (living or non-living) (29), this assay was a reliable measure of the isolate’s capacity to adhere to abiotic surfaces. Isolates classified as non-biofilm producers under the biofilm assay were regarded as having a weak adhesion ability, those regarded as weak to moderate biofilm producers were classified to have moderate adhesion capacity, and those with strong to extremely strong biofilm were reclassified as having strong abiotic adhesion ability.

#### 2.6.5. Phospholipase Activity

Phospholipases are hydrolytic enzymes involved in degradation of lipid constituents of host cell membranes, thereby promoting tissue invasion and damage (27,35). Phospholipase activity was determined according to the test procedure discussed in other previous studies, with some modifications (30,36). A volume of 5 μL of standard yeast suspension from fresh culture was inoculated as a spot-on SDA supplemented with 10% fresh egg yolk emulsion. The plates were inoculated in duplicates and incubated for up to 72 hours. The diameters of the colonies and the halo or precipitation formed around them were measured. The phospholipase zone (Pz) was determined by dividing the colony diameter by the precipitation zone plus colony diameter. The isolates in our study were classified as follows: Pz = 1, negative phospholipase activity; 0.8 to 0.98 as weak; 0.65 to 0.79 as moderate; below 0.64 as strong phospholipase producers.

#### 2.6.6. Haemolysin activity

Haemolysins are hydrolytic enzymes that lyse host red blood cells to release iron, a crucial micronutrient for fungal growth and proliferation (27). Haemolysin activity was tested on SDA supplemented with blood as per previous studies reported elsewhere (30,36). A volume of 5 μL standard yeast suspension from fresh culture was inoculated as a spot-on SDA supplemented with 10% human blood and 3% glucose in duplicates and incubated for up to 48 hours in a CO_2_-elevated atmosphere. The presence of a clear halo around the inoculum indicated positive haemolysis. The diameter of colonies and zones of haemolysis was measured to obtain the haemolysis index (HI) for each isolate. HI was determined by dividing the colony diameter by the haemolysis zone plus colony diameter. The isolates in our study were classified as follows: HI = 1, negative haemolysin activity; 0.76 to 0.98 as weak; 0.61 to 0.75 as moderate; below 0.60 as strong haemolysin producers.

#### 2.6.7. Catalase Activity

The role of catalases is to break down hydrogen peroxide into water and oxygen (37,38). Hydrogen peroxide is produced by certain white blood cells, especially neutrophils, mainly to kill pathogens (38). Catalase activity was tested using the hydrogen peroxide method (39). A drop of 3% hydrogen peroxide (H_2_O_2_) was placed on an agglutination tile. Using a sterile inoculation loop, the test organism from fresh cultures was emulsified onto the drop of H_2_O_2_. Immediate bubbling indicated catalase-producing yeast species, while non-catalase-producing organisms did not produce bubbles upon contact with the H_2_O_2_. The bubbling was graded based on the amount, time to bubble, and time to clear out. Therefore, based on this, the isolates were graded as strong catalase producers, moderate producers, weak producers, and non-catalase producers.

#### 2.6.8. Urease Activity

Under nutrient-deficient environments, urease often plays a role in the nitrogen assimilation for pathogenic microbes and raises pH to allow colonisation and survival (84,85,86). Urease activity was determined using Urea Agar media (Himedia Laboratories, Mumbai, India) as in other previous studies (41,42). The inoculated Urea Agar test tubes were incubated for up to 48 hours. The colour changes of the media from yellow/orange to pink indicated urease activity. Colour change was quantified macroscopically, taking note of the intensity of the pink colour and the completeness of the change of the media colour. Therefore, the isolates were classified based on this as strong urease producers, moderate producers, weak producers, and non-urease producers.

### 2.7. Data Analysis

Data was analysed in Microsoft Excel 2016 and R Studio (Version 4.5.2, 2025.09.2) using descriptive statistics to determine the frequency of yeast species and their virulence factors that were presented in tables and graphs. To determine similarities in enzymatic activities among the species, a hierarchical clustering heat map was generated with Excel. ArcGIS Pro (Version 3.1) was utilised to create a map showing sampling points of the environmental samples and the distribution of environmental yeast species within the study area.

## 3.0. Results

### 3.1. Fungal Diversity and Distribution

Of the 71 environmental yeast isolates recovered based on distinct colony morphology on SDA, 54 (75%) were identified. From each sample, isolates recovered at different temperatures but identified as the same species were grouped, and only the thermotolerant isolates were analysed further. This yielded 33 distinct environmental isolates. The clinical and environmental isolates spanned five genera: *Candida*, *Pichia*, *Komadaea*, *Trichosporon*, and *Nakaseomyces*. The 33 environmental isolates comprised 39% (n=13) *Pichia kudriavzevii (*formerly *Candida krusei)*, 30% (n=10) *Candida orthopsilosis (Candida parapsilosis* group II), 6% (n=2); *Candida tropicalis*, *Candida guilliermondii,* and *Candida lambica,* 3% (n=1); *Nakaseomyces glabrata (*formerly *Candida glabrata), Candida albicans, Komadaea ohmeri,* and *Trichosporon asahii.* The clinical isolates comprised 29% (n=6) *Candida Parapsilosis sensu stricto,* 24% (n=5) *C. albicans,* 10% (n=2); *N. glabrata, P. kudriavzevii,* and *C. tropicalis,* 5% (n=1); *Candidozyma auris (*formerly *Candida auris), Candida rugosa, K. ohmeri,* and *T. asahii* (Table 1). Among the 33 environmental isolates, 82% (n=27) belong to fungal species listed within the WHO fungal pathogen priority groups, while 86% (n=18) of the clinical isolates belong to fungal species listed within the WHO fungal pathogen priority groups.

**Table 1:**
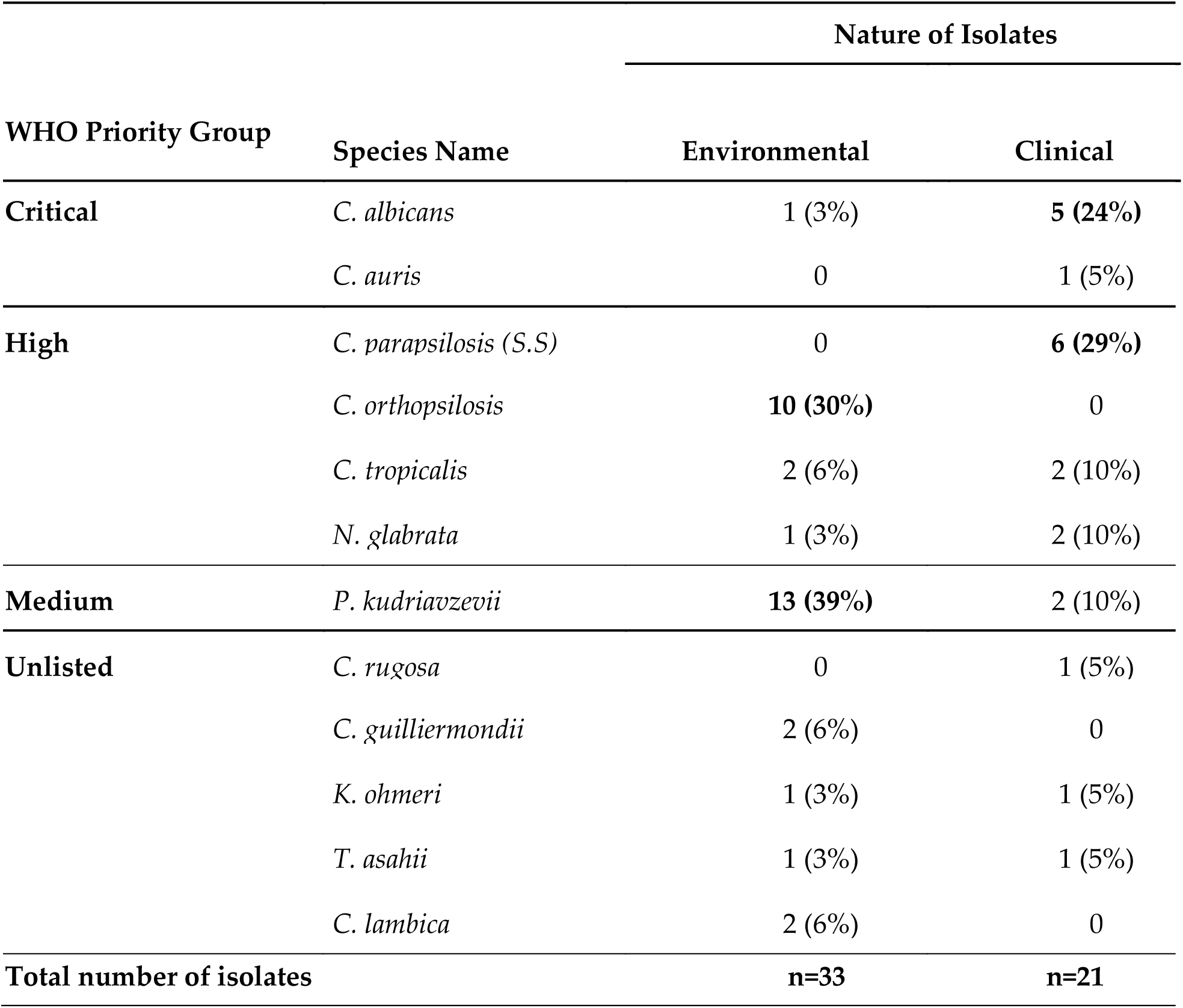
Yeast species from guano-contaminated environments and clinical data and their respective ranks on the WFPPL. The two data sets are dominated by fungal species belonging to critical and high-priority groups.

| WHO Priority Group | Species Name | Nature of Isolates |  |
| --- | --- | --- | --- |
|  |  | Environmental | Clinical |
| **Critical** | *C. albicans* | 1 (3%) | **5 (24%)** |
|  | *C. auris* | 0 | 1 (5%) |
| **High** | *C. parapsilosis* (S.S) | 0 | **6 (29%)** |
|  | *C. orthopsilosis* | **10 (30%)** | 0 |
|  | *C. tropicalis* | 2 (6%) | 2 (10%) |
|  | *N. glabrata* | 1 (3%) | 2 (10%) |
| **Medium** | *P. kudriavzevii* | **13 (39%)** | 2 (10%) |
| **Unlisted** | *C. rugosa* | 0 | 1 (5%) |
|  | *C. guilliermondii* | 2 (6%) | 0 |
|  | *K. ohmeri* | 1 (3%) | 1 (5%) |
|  | *T. asahii* | 1 (3%) | 1 (5%) |
|  | *C. lambica* | 2 (6%) | 0 |
| **Total number of isolates** |  | **n=33** | **n=21** |

From each of the sampling locations, at least 4 fungal species were isolated. *Pichia kudriavzevii* and *C. orthopsilosis* were isolated from all the peri-urban areas under the study area. Certain species, were only isolated from specific locations across the entire study area. For instance, *N. glabratus* and *C. tropicalis* were only recovered in samples collected in the Mbayani-Chemusa area, *C. guilliermondii, T. asahii,* and *C. albicans* were only isolated from samples from the Ndirande area, while *C. lambica* and *K. ohmeri* were only isolated from samples collected in the Chilobwe area (Figure 1).

**Figure 1:**
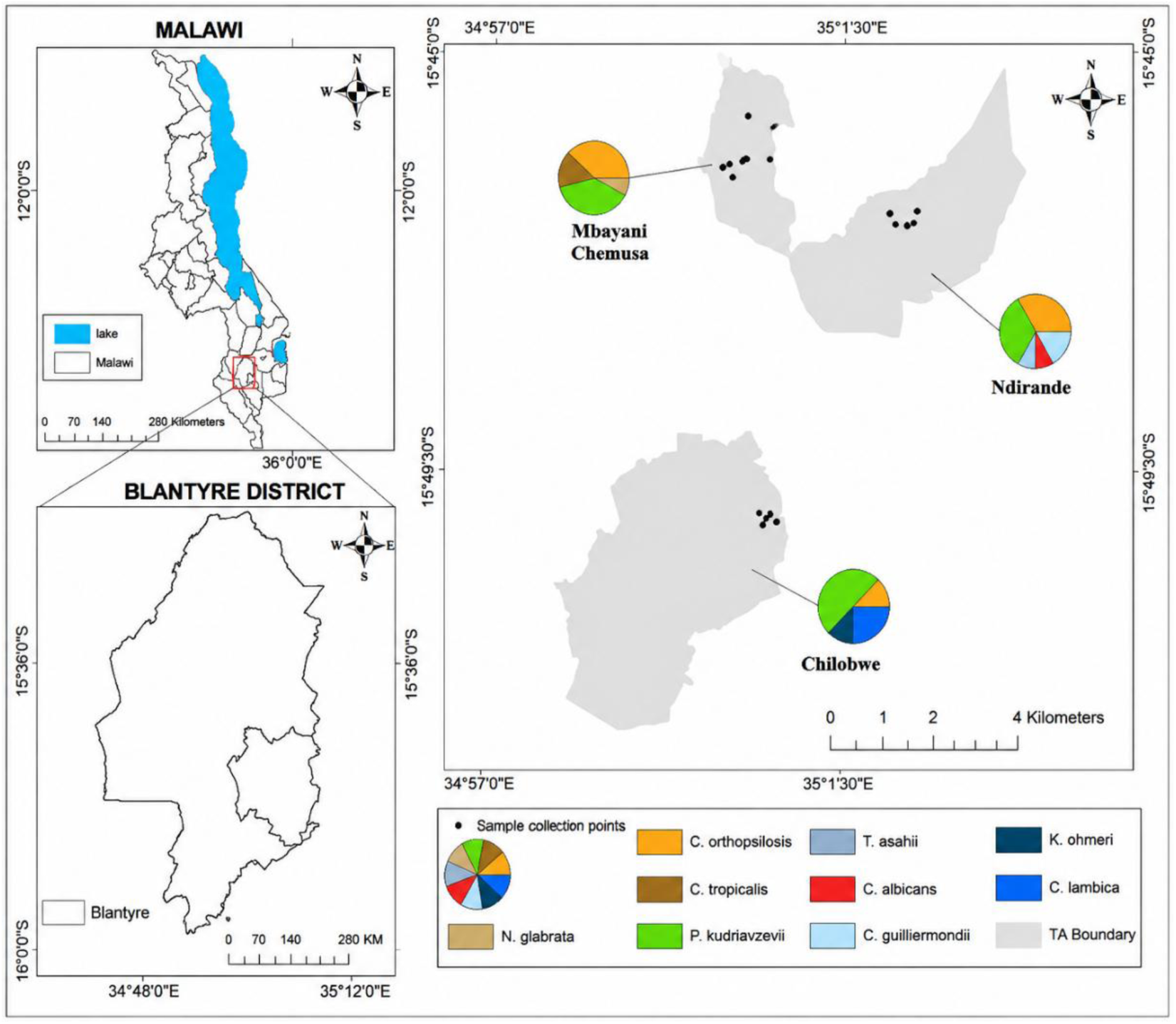
Guano collection points within each peri-urban area in Blantyre, Malawi and the distribution of the medically important yeast species in the study area. *Pichia kudriavzevii* and *C. orthopsilosis* are present in all peri-urban areas.

Fifty per cent (50%) of the species identified from the environmental samples were also identified among the clinical isolates. *Candidozyma auris*, *C. parapsilosis sensu stricto*, and *C. rugosa* were only observed among the clinical fungal isolates, while *C. orthopsilosis*, *C. guilliermondii*, and *C. lambica* were only observed among the environmental isolates recovered from the pigeon guano contaminated environments (Figure 2).

**Figure 2:**
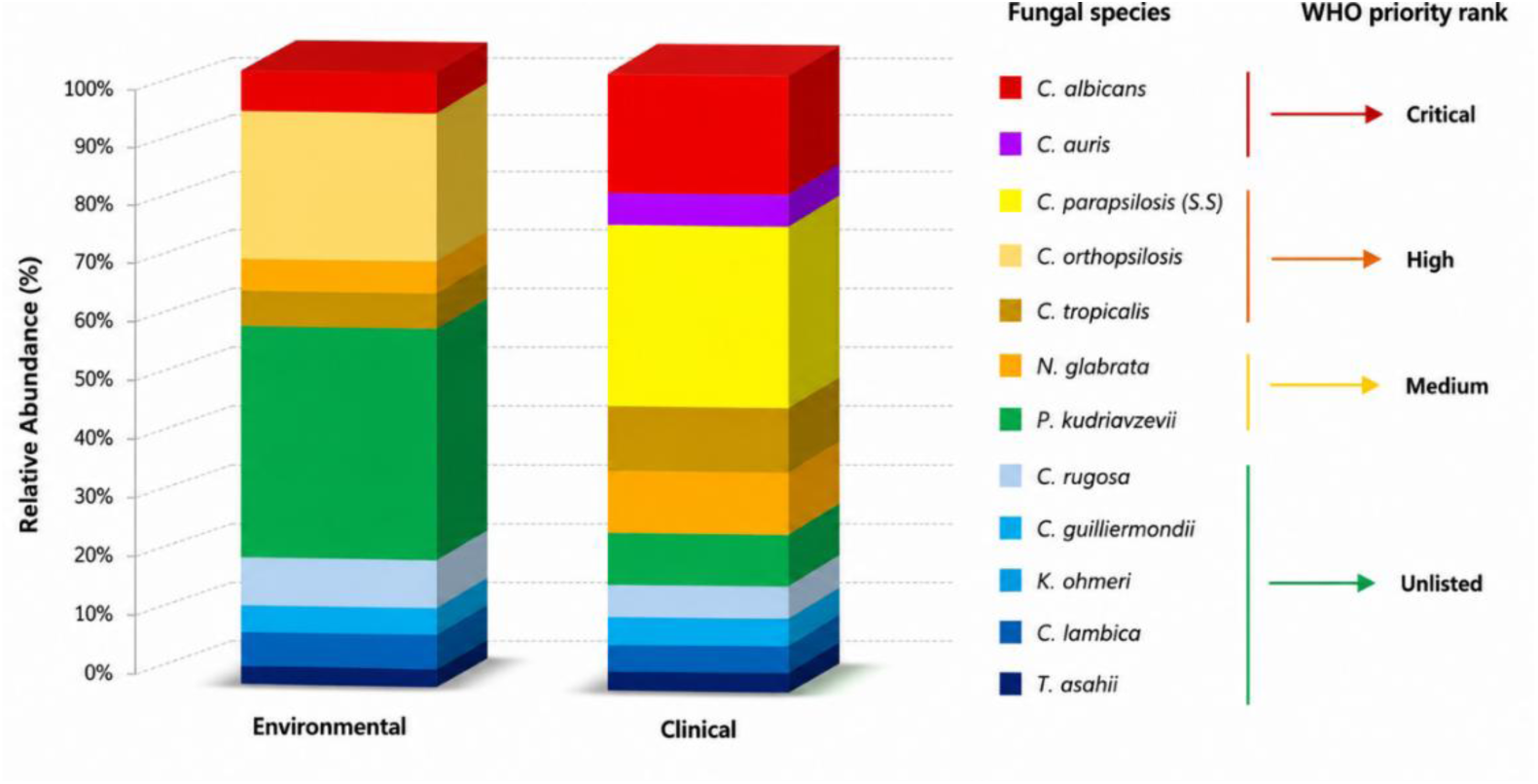
Overlap in fungal species composition between clinical data and environmental fungal isolates: 50% of the taxa are shared

### 3.2. Virulence Factors

A majority of the environmental and clinical isolates showed a positive reaction to the virulence factors tested. Results from the virulence tests and how a reaction was categorised as negative, weakly positive or strongly positive are shown in Figure 3.

**Figure 3:**
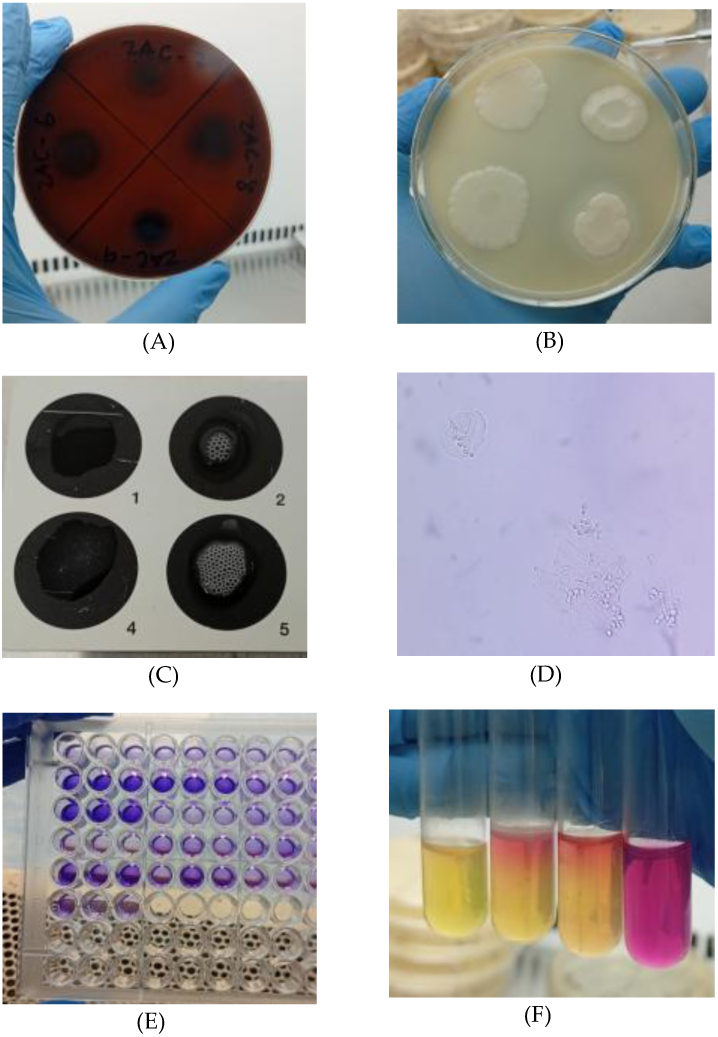
Visual plate depicting positive and negative responses to virulence-factor assays across environmental and clinical yeast isolates. **(A). Haemolysin activity;** note a dark spot where the yeast suspension was inoculated, and the zone around it. Some colonies have two zones around the yeast colony: a darker zone and another clear zone around the darker zone. Colonies with double zones are an indication of more than one haemolysin activity **(B). Phospholipase activity**; note that the colonies look unusually big on SDA supplemented with egg yolk emulsion, with some colonies having an opaque zone around the spot of inoculation, which looks hollow. The opaque zone is an indication of strong phospholipase activity. **(C). Catalase activity:** visible bubbling after mixing a yeast colony with 3% hydrogen peroxide. Spot 4 has small bubbles, and this was classified as a weak catalase reaction, while spot 5 has big bubbles, and this was classified as a strong catalase reaction. **(D). Adhesion to biotic surface;** blastoconidia are seen in this plate, adhered to buccal epithelial cells. **(E). Biofilm formation;** destaining solution in a new polystyrene microplate after solubilisation of bound crystal violet in the well with absolute ethanol. Note that some wells look sharp blue as an indication of strong biofilm formation, and some wells look light blue as an indication of weak biofilm formation. (**F). Urease activity;** note the colour change in the urea agar media from orange to pink. The test tube at the far right shows a negative urease reaction. The two test tubes in the middle show only colour change at the top half of the tube, an indication of weak urease activity and the test tube on the left side is completely pink, an indication of strong urease activity.

#### 3.2.1. Thermotolerance

Since all clinical isolates were obtained from human samples, they were all regarded as thermotolerant. Eighty-two per cent (n=27) of the environmental isolates were able to grow only at 37°C, while 18% (n=6) only showed growth at room temperature; therefore, they were regarded as thermosensitive. All 6 thermosensitive isolates were *Candida orthopsilosis*.

#### 3.2.2. Biofilm Formation

We observed varied OD_570_ nm values from all the study isolates, results evident from the solubilised biofilms in the microplates (Figure 3E). For environmental isolates, the OD_570_ nm values ranged from 0.28 to 3.96, with a median value of 0.86. Positive biofilm formation (OD_570_ nm > 0.83) was observed in 13 (39%) of the isolates. Of these isolates, 7 (21%) exhibited weak biofilm formation, while 6 (18%) isolates, *C. orthopsilosis*, *C. tropicalis*, and *T. asahii,* exhibited moderate biofilm formation. For the clinical isolates, OD_570_ nm values ranged from 0.32 to 5.51, with a median value of 0.82. Only 6 (29%) of the isolates were positive for biofilm, with 4 (19%) exhibiting weak biofilm values, 1 (5%) exhibiting moderate biofilm values, and the remaining 5% also exhibiting strong biofilm values. The isolates that exhibited moderate and strong biofilm values were *T. asahii* and *C. parapsilosis,* respectively (Table 2).

**Table 2.**
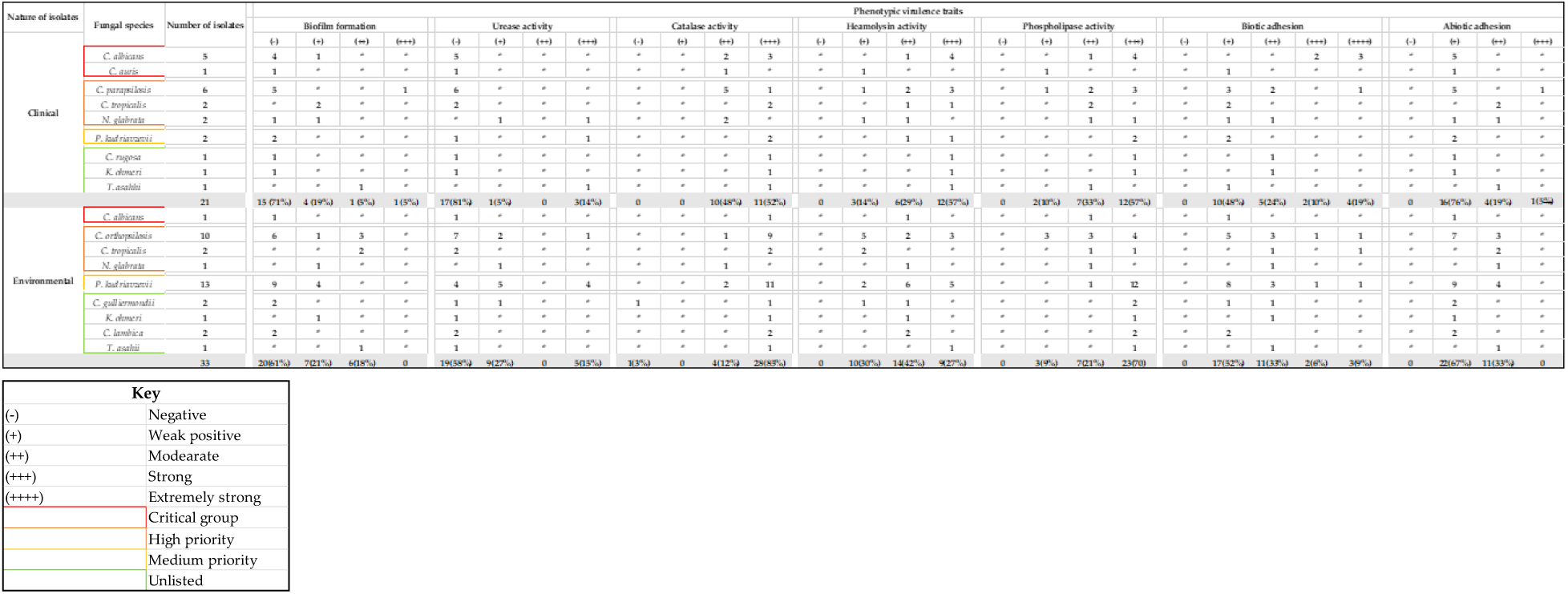
Species-level virulence-factor scores across clinical and environmental isolates, with counts and percentages for positive and negative outcomes.

#### 3.2.3. Adhesion to Biotic Surface

All isolates showed the ability to adhere to HBEC (Figure 3D). Among environmental isolates, the number of blastoconidia attached to 50 HBEC ranged from 2 to 94 blastoconidia, with a median of 10 cells per 50 HBEC. Of the 33 environmental isolates, 17 (52%) exhibited weak adhesion ability, 11 (33%) showed moderate adhesion ability, and 2 (6%); *P. kudriavzevii* and *C. orthopsilosis* showed strong adherence, and 3 (9%) isolates comprising *P. kudriavzevii*, *C. orthopsilosis*, and *C. tropicalis* showed extremely strong adherence to the HBEC. Among the clinical isolates, the number of adhered blastoconidia ranged from 4 to 67 cells per 50 HBEC, with a median of 13 cells per 50 HBEC. Out of the 21 isolates, 10 (48%) isolates showed weak adherence, 5 (24%) showed moderate adherence, 2 (9%) isolates, all *C. albicans,* showed strong adhesion ability, and 4 (19%) isolates comprising *C. albicans* and *C. parapsilosis* showed extremely strong adherence to the HBEC (Table 2).

#### 3.2.4. Adhesion to Abiotic Surface

Twenty-two (67%) environmental isolates were classified as having a weak abiotic surface adhesion ability, while moderate adherence was observed in 11 (33%) isolates: *P. kudriavzevii*, *C. orthopsilosis*, *C. tropicalis*, *N. glabrata*, and *T. asahii*. We did not observe any isolates having a strong adherence to the abiotic surface. Regarding the abiotic adherence ability in clinical isolates, 16 (76%) isolates showed a weak ability, moderate adherence was observed in 4 (19%) isolates: *C. tropicalis*, *C. albicans*, *N. glabrata*, and *T. asahii*. Strong adherence was observed only in 1 isolate of *C. parapsilosis* (Table 2).

#### 3.2.5. Phospholipase Activity

All isolates showed positive phospholipase activity, Pz < 1 (Figure 3B). For environmental isolates, the Pz values ranged from 0.42 to 0.93 with a mean value of 0.62. Of the 33 environmental isolates, 3 (9%) isolates exhibited weak phospholipase activity, and moderate phospholipase activity was observed in 7 (21%) isolates. Strong phospholipase activity was observed in 23 (70%) isolates, with a majority being *P. kudriavzevii* and *C. orthopsilosis*. In terms of the clinical isolates, the Pz values ranged from 0.30 to 0.83 with a mean value of 0.63. Of the 21 isolates, 2 (10%) isolates exhibited weak phospholipase activity, 7 (33%) showed moderate phospholipase activity, and 12 (57%) isolates showed strong phospholipase activity, especially *C. albicans*, *C. parapsilosis*, and *P. kudriavzevii* (Table 2).

#### 3.2.6. Haemolysin Activity

All isolates showed positive haemolysin activity, HI < 1 (Figure 3A). The HI values among the environmental isolates ranged from 0.38 to 0.91, with a median value of 0.73. Of the 33 environmental isolates, 10 (30%) exhibited a weak haemolysin activity, moderate haemolysin activity was observed in 14 (42%) isolates, while strong haemolysin activity was observed in 9 (27%) isolates, with a majority again being *P. kudriavzevii* and *C. orthopsilosis*. Regarding the clinical isolates, the HI values ranged from 0.33 to 0.89, with a median value of 0.59. Out of the 21 isolates, 3 (14%) isolates showed a weak haemolysin activity, 6 (29%) showed moderate haemolysin activity, and 12 (57%) showed strong haemolysin activity, mainly *C. albicans* and *C. parapsilosis* (Table 2).

#### 3.2.7. Urease Activity

Urease activity was indicated by a colour change in the urea agar medium from orange to pink (Figure 3F). Out of the 54 isolates tested for urease activity, only 18 (33%) showed a positive urease activity. Among the 18 positive isolates, 14 (78%) were environmental isolates consisting of *P. kudriavzevii*, *C. orthopsilosis*, *N. glabrata*, and *C. guilliermondii*. Only 4 clinical isolates tested positive for urease, including *P. kudriavzevii*, *N. glabrata*, and *T. asahii* (Table 2).

#### 3.2.8. Catalase Activity

All 54 isolates showed positive catalase activity, but varied in the degree of activity (Figure 3C). Among the environmental isolates, strong catalase activity was observed in 28 (85%) isolates, moderate catalase activity was observed in 4 (12%) isolates, and weak catalase activity was only observed in *C. guilliermondii*. Among the clinical isolates, a strong catalase reaction was observed in 11 (52%) isolates, and the remaining 10 (48%) isolates exhibited moderate catalase activity (Table 2).

Overall, the majority of the environmental isolates showed the ability to produce more than one key virulence factor, traits that were similar to those of the clinical isolates. The commonly observed virulence factors in both the environmental and clinical isolates were catalase, phospholipase, and haemolysin. The virulence factor traits between critical species, high-priority species and medium-priority species broadly mirrored (Figure 3)

## 4.0 Discussion

### 4.1 Fungal Diversity and Distribution

To our knowledge, this is the first study in Malawi isolating medically important yeast species in environments contaminated with guano of synanthropic pigeons and comparing their characteristics with clinical yeast species. All the environmental isolates in the current study were of clinical significance. The majority (52%) belonged to the genus *Candida,* findings consistent with other studies (10,25,43–46). Nevertheless, other investigations have reported dominance by non-*Candida* taxa, particularly *Cryptococcus* species (15). Interestingly, the clinical isolates in the current study were even more dominated (71%) by *Candida* species, a pattern that also mirrors results from studies conducted elsewhere (47–49) and is reflective of the predominance of *Candida* species on the WHO fungal priority pathogen list (12). Even though *Candida* spp. forms part of the microbiota of healthy humans and animals, many species are associated with nosocomial bloodstream infections, invasive candidiasis, and superficial mycoses (45,50). Acquisition is either endogenous or exogenous, facilitated by environmental sources that introduce contaminations (45).

*Pichia kudriavzevii* (39%) and *C. orthopsilosis* (30%) constituted a majority of the environmental isolates (Table 1), patterns similar to reports elsewhere (25,51). *Pichia kudriavzevii* is increasingly recognised as an important cause of nosocomial outbreaks (52,53), and is a well-established pathogen due to inherent resistance to fluconazole and reduced susceptibility to amphotericin B (54). Several other studies have reported findings dominated by *C. guilliermondii*, *C. albicans*, and *N. glabrata* (10,45,55–57). Geographical range, even within a region/nation, may contribute to variations in the epidemiology of microorganisms like fungi (27,58). However, *P. kudriavzevii* and *C. orthopsilosis* were isolated in all the peri-urban areas in our study (Figure 1). On the other hand, clinical yeast isolates were dominated by *Candida parapsilosis sensu stricto* (29%) and *C. albicans* (24%), findings consistent with a 20-year sentry surveillance program (1997–2016) that reported a progressive increase in *C. parapsilosis* and *N. glabrata* cases and a concurrent decrease in *C. albicans* cases across the years (59). Although *P. kudriavzevii* dominated the environmental sample set in our study, it was less well represented among the clinical isolates (10%). Interestingly, *P. kudriavzevii* is very commonly implicated in clinical cases in North America, Europe, and some parts of Asia (60).

We noted that environmental isolates and clinical isolates shared 50% of the species in common in the current study (Figure 2). This finding aligns with a growing body of literature showing substantial overlap between yeasts recovered from avian-environmental sources and clinical cases (10,15,25,27,57). This overlap may suggest potential environmental reservoirs contributing to human exposure and, under appropriate conditions, to infection. Other species like *C. albicans* (3%), *N. glabrata* (3%), and *C tropicalis* (6%) were relatively low in the environmental samples (Table 1), but they are highly ranked on the WHO fungal pathogen priority list. *Komadae ohmeri* and *T. asahii* are not listed as a priority by WHO. These species are usually reported at comparatively low levels in both clinical and environmental settings (10,15,25,27,57), but are medically important because of their invasive nature and mortality rates that range between 50% and 80% (61,62), which surpass some of the notable species on the WHO list (12).

### 4.2 Virulence Factors

The majority (82%) of the environmental isolates in our study were able to grow at 37°C. Consistently, other studies have reported significant thermotolerance in environmental yeast (10,15,30,45,63–65). Only some strains of *C. orthopsilosis* showed a sense of thermosensitivity. Nevertheless, *C. orthopsilosis* has been implicated in several cases, especially in neonates (66), and is also emerging due to selective pressure from rising temperatures (67).

Although no environmental isolate in our study was regarded as a strong biofilm producer, 50% of the isolates showed moderate biofilm-forming ability. In contrast, only a single clinical isolate showed moderate to strong biofilm-forming ability, and the rest had none (Table 2). This is likely an adaptation to the harsher environmental conditions compared to those of a human host. For instance, we recovered two isolates of *C. tropicalis* from environmental samples and another two from clinical samples. All the *C. tropicalis* isolates from the environment produced significantly higher biofilm values than the two clinical isolates. Similar findings have also been reported elsewhere (30,35). It has been widely reported that the process of biofilm formation varies among species and depends on where they have been isolated (29,68). *Trichosporon. Asahii* and *C. parapsilosis* species complex showed a stronger ability to form biofilms in both clinical and environmental isolates. The link between adhesion to abiotic surfaces and biofilm formation implies that species such as *T. asahii, C. parapsilosis* species complex, and *C. tropicalis* are more likely to adhere to abiotic surfaces like cannulas, catheters, tubes, and some artificial devices. Comparatively stronger adhesion by *C. parapsilosis* and *C. tropicalis* to such surfaces than the other species has been reported (29,68).

All isolates in our study demonstrated the ability to adhere to the HBEC (biotic adhesion). Notably, clinical isolates (29%) adhered more strongly to the HBEC than environmental isolates (15%). By contrast, others have previously reported environmental isolates with a stronger adherence to HBEC than clinical isolates (30). Adhesion is influenced by species-specific differences, as the in vitro adhesion process varies among fungal species (29). Among environmental species, *P. kudriavzevii*, *C. orthopsilosis*, and *C. tropicalis* consistently exhibit strong adhesion properties, aligning with findings from other reports (30,34). These are thus more likely to colonise or initiate an infection if they find their way into a human host. *C. albicans* has been described as the most adherent species to epithelial cells in studies focusing on clinical isolates (29,69), a pattern that our clinical data also reflect (Figure 4, Table 2).

**Figure 4:**
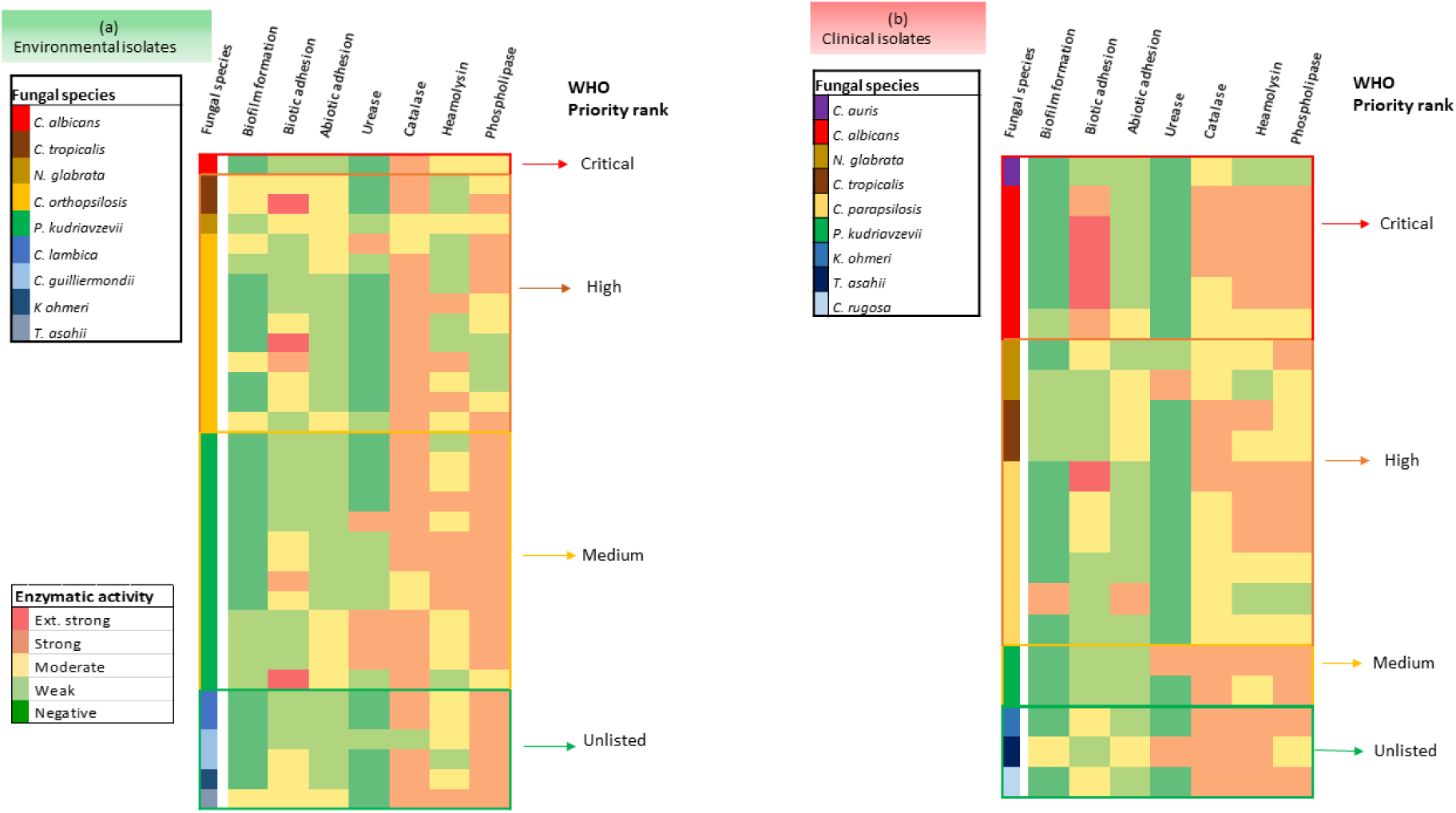
Virulence-factor matrix across environmental and clinical isolates; species profiles are broadly similar.

A majority of the environmental isolates (70%) in the current study showed strong phospholipase activity, relative to clinical isolates (57%). Similar findings have been documented elsewhere (70), while other studies have shown no phospholipase activity in both clinical and environmental isolates (35). Strain-specific differences may account for these conflicting observations. Strong phospholipase activity in the environmental isolates was commonly observed in *P. kudriavzevii* and *C. orthopsilosis*. *Candida albicans* and *C. parapsilosis* were the common species exhibiting strong phospholipase activity among the clinical isolates. These associations likely reflect the species composition of the isolates tested in this study; environmental samples were dominated by *P. kudriavzevii* and *C. orthopsilosis*, while clinical samples were dominated by *C. albicans* and *C. parapsilosis* (Table 1). It is possible that testing a broader or different set of isolates could yield different patterns of enzyme activity, a pattern seen in other studies (71–73).

All isolates in the current study showed haemolytic activity. However, strong haemolytic activity was observed mainly in clinical isolates (57%) and only in 27% of the environmental isolates, trends also reported elsewhere (30). All clinical isolates showed an indication of multiple haemolysins, indicative of their adaptation to the human host, where iron is very restricted. *Candida* species in particular often show strong haemolytic activity (27,36,74,75), as was observed in the current study. Strong haemolysin activity shown by some species like *P. kudriavzevii*, *C. orthopsilosis*, and *T. asahii* in this study could also explain why these species are increasingly becoming an important cause of nosocomial infections (52,62,66), while similar haemolytic trends in some species have also been reported elsewhere (76).

Catalase activity was stronger in environmental isolates (85%) than in clinical isolates (52%). As it is known that catalase-deficient strains are more vulnerable to oxidative stress caused by immune attacks (38), our findings could be more related to environmental stress. Although the link between environmental stress and virulence is still being extensively researched, stressed microorganisms tend to be more virulent (80,81). Similar trends have also been reported elsewhere (37,41).

Among all the environmental isolates, positive urease activity was observed in 42% and only 19% in the clinical isolates. Our findings are consistent with reports that most yeast species responsible for causing infections in humans are usually urease negative and that only a few environmental strains show urease activity (77,78). Nevertheless, others have reported urease activity in more than 50% of other environmental yeast species (41). In the current study, environmental isolates were dominated by *P. kudriavzevii*, a species mostly reported as a urease producer (79). Just like catalases, urease-deficient strains often show weak virulence and are more susceptible to macrophage-mediated killing due to their inability to prevent the acidification of phagosomes (40).

### 5.0 Conclusion

This study provides important data regarding the diversity, distribution, and pathogenic potential of medically important yeast species in pigeon-guano-contaminated environments within the built environment in Blantyre, Malawi. A majority of the isolates from the guano-contaminated environments are listed as priority by the World Health Organization and also showed phenotypic traits of virulence similar to clinical yeast isolates, meaning that they are pre-adapted for invasion, making it a public health concern. Our shared ecosystems may facilitate the movement of these potentially pathogenic fungi between pigeons, human-habituated environments, and humans or other animals. The high prevalence and distribution pattern of *P. kudriavzevii* is worrisome, considering that they are naturally resistant to some azoles, microorganisms can share resistance genes in the environment, and pigeons can cover long distances, increasing the potential risk of resistant pathogen spread. This study also reports a rise in *non-albican* species in the human population, with most of the species also circulating in pigeon guano-contaminated environments. These findings call for the critical need for integrated fungal pathogen surveillance and targeted public health interventions in urban ecosystems where birds and their environments pose an actionable risk to vulnerable populations.

## Declarations

### Author Contributions

Conceptualisation, B.M.; P.C.; G.B.; methodology, B.M.; P.A.; P.C.; G.B.; formal analysis, B.M.; P.A.; resources, B.M.; P.C.; G.B.; data curation, writing-original draft preparation, B.M.; writing-review and editing, B.M.; P.A.; P.C.; G.B.; supervision, P.C.; G.B.; funding, B.M. All authors have read and agreed to the published version of the manuscript.

### Clinical Trial Number

Not applicable.

### Ethical Approval and Consent to Participate

The study was reviewed and approved by the Malawi University of Science and Technology Research Ethics Committee (Reference No. P.12/2024/373). The research complied with the Helsinki Declaration. The acquisition of clinical isolates did not involve handling humans or human-derived samples; only isolates that had previously been collected from patients during routine clinical analyses by laboratory personnel were used, and the isolates were collected without any identifiable patient information. The clinical isolates were assigned sample IDs to ensure privacy and confidentiality. Approval to use the clinical fungal isolates was granted by the laboratories responsible for each facility. Before study initiation, we obtained study-site authorisation from Blantyre District Council. All participants/household owners involved in this study gave their formal informed consent verbally before collecting pigeon guano samples from their households.

### Consent for Publication

Not applicable

### Availability of Data and Materials

The original contributions presented in this study are included in the article.

## Data Availability

All data produced in the present work are contained in the manuscript

## Acknowledgements

We would like to thank the NORHED II One Health Project at the Malawi University of Science and Technology for the scholarship opportunity. The authors also extend their sincere gratitude to the household owners in the study area for their approval to collect samples. We are deeply indebted to Mr R. Kondwani, the research assistant, who tirelessly assisted with the identification of households that keep pigeons, seeking initial approval, and assisting with sample collection. We acknowledge the support and cooperation of the Blantyre District Council, the Malawi Liverpool Welcome Programme, the Zomba Central Hospital Microbiology Laboratory, the Public Health Institute of Malawi Microbiology Laboratory, and the University of Malawi. Special thanks are extended to Mr Harvey Chilembwe, Mr Vitumbiko Chijere Chirwa, Mr Dyson Kwenda, and the MUST One Health Cohort 2 members for their contribution to this study.

